# Weight-Related Hypertension in Youth: Evaluation of Family Nutrition and Physical Activity Tool

**DOI:** 10.1101/2024.08.21.24312365

**Authors:** Lisa Bailey-Davis, Carolyn F. McCabe, Chengpeng Zeng, Karissa L. Peyer, Samuel S. Gidding, Adam Cook, G. Craig Wood, Jennifer Franceschelli Hosterman, Shuping Ge, Gregory J. Welk

**Author notes:** Address correspondence to: Lisa Bailey-Davis, Department of Population Health Sciences and Center for Obesity & Metabolic Research, Geisinger. 100 N Academy Ave, Danville, PA. Contributed equally as co-first authors (Lisa Bailey-Davis, Carolyn F. McCabe). **Funding/Support**: No funding was secured for this study. **Abbreviations:** Family Nutrition and Physical Activity (FNPA); American Academy of Pediatrics (AAP); U.S. Preventive Service Task Force (USPSTF); Body Mass Index (BMI); Hypertension (HTN); Hazard ratio (HR); Confidence interval (CI).

## Abstract

**Objectives and Background:** This study explores the potential moderating influence of Family Nutrition and Physical Activity (FNPA) behavior scores on reducing hypertension incidence associated with overweight changes during childhood.

**Methods:** A longitudinal design was used to explore associations between children’s FNPA scores and overweight status change (when ages 3-9 years) on hypertension during latter childhood and adolescence (ages 9-18 years). Data were obtained from well-child visits. Participants were classified into 3 by 3 subgroups by FNPA score (low, moderate, high) and overweight status (loss, stable, gain). Cox proportional hazards models were used to estimate hazard ratios of hypertension development with the moderate FNPA score X overweight stable reference group.

**Results:** Among 3808 participants, 58% were publicly insured. Among participants who gained overweight during childhood, 41.7% developed hypertension compared to 29.9% who were overweight stable and 27.3% who lost overweight (P <0.001). After accounting for confounders, participants who gained vs. maintained overweight had 2.01 (95% CI 1.65-2.44) times higher hypertension risk. Although FNPA scores were not associated with hypertension incidence, the interaction term between a high FNPA and overweight gain was significant (P = 0.01). Among children who gained, hazard ratios for hypertension among low, moderate, and high FNPA groups were estimated at 1.99 (95% CI 0.98-4.05), 2.01 (95% CI 1.64-2.44), 1.54 (95% CI 0.71-3.34), respectively.

**Conclusions:** FNPA screening can inform preventive counseling about healthy home environments. Behaviors associated with high FNPA scores potentially reduce hypertension risk among children who experience overweight gain by approximately 25%.

**Article Summary:** Having higher, healthier Family Nutrition and Physical Activity behavior scores moderates the incidence of hypertension associated with gain in overweight during early-middle childhood.

**What’s Known on This Subject:** The AAP Clinical Practice Guidelines for the Screening and Management of High Blood Pressure provides recommendations which naturally accord with those for obesity prevention and management. The FNPA has been used in clinical settings to facilitate implementation of anticipatory guidance.

**What This Study Adds:** The results underscore the novelty and importance of FNPA as a screening tool and in identification of the positive culmination of behaviors, reflected as high FNPA score, in moderating the risk of hypertension among children experiencing gain in overweight status.

## Introduction

Many chronic conditions plaguing public health are linked to obesity, and when established early in life, obesity is associated with adverse cardio-kidney-metabolic outcomes (e.g., high blood pressure, abnormal lipid levels, chronic kidney disease, and insulin resistance). In a meta-analysis of studies across the globe, obesity was positively associated with the increasing prevalence of childhood hypertension observed in the last two decades.^1^ During childhood, hypertension tracks with increasing body mass index, with prevalence rates as high as 14% among children with overweight^2^ and 25% among those with obesity.^3^ Substantial early morbidity is associated with childhood hypertension that, like obesity, persists into adulthood and results in potentially irreversible cardio-kidney-metabolic organ damage.^4,5^ Given that the prevalence of childhood hypertension is directly linked to obesity, upstream solutions are needed to prevent obesity.^6^

Pediatricians promote healthy home environments and behaviors to prevent obesity and associated downstream conditions such as hypertension. The American Academy of Pediatrics Clinical Practice Guidelines for the evaluation and treatment of children and adolescents with obesity recommend pediatricians screen children for overweight and obesity beginning at age 2 years and assess family and home environment factors related to nutrition and physical activity behaviors to inform preventive counseling.^7^ Additionally, the U.S. Preventive Service Task Force (USPSTF) recommends (grade B) screening for obesity in children and adolescents aged 6 years and older.^8^ Screening includes assessing risk related to poor nutrition, low levels of physical activity, inadequate sleep, and sedentary behaviors. The Family Nutrition and Physical Activity (FNPA) behavior screening is a validated tool that was developed to capture home environments and practices that influence these obesogenic behaviors. Family-based practices and youth behaviors are assessed by the parent, with summary scores ranging from 20 to 80 (Table 1).^9^ Prior studies have consistently observed an inverse pattern of association between FNPA summary scores and obesity risk, e.g., lower FNPA summary scores are associated with greater odds of having or developing obesity and higher scores are associated with a healthier weight category.^10–14^ The FNPA has been adopted by health systems as a risk assessment tool to enhance prevention / treatment and has been used to guide diverse family groups, at different life stages, living in varied socioeconomic conditions, and facing co-morbidities that exacerbate obesity risk.^13–16^ Including FNPA in standard well-child visits helps prevent childhood obesity.^13^ However, studies to date have not yet examined the utility of the FNPA for predicting other related clinical outcomes.

**Table 1.**
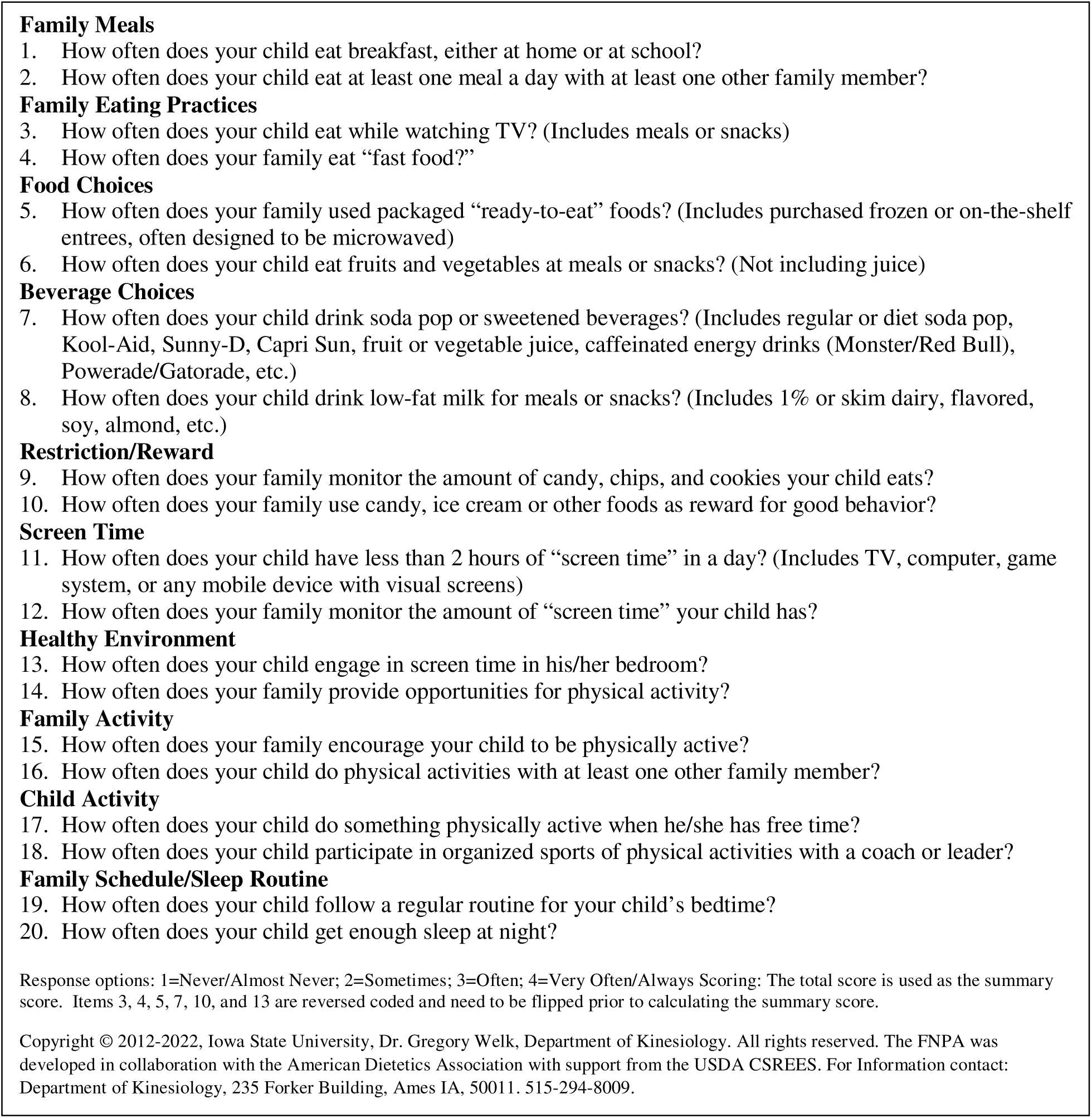
Family Nutrition and Physical Activity Screening Tool.

This retrospective, observational cohort study from a large health system used longitudinal data obtained from clinical well-child visits to evaluate the interaction of weight gain among children with overweight and FNPA risk score on incident hypertension. Our secondary purpose was to explore the potential moderating influence of FNPA as a global measure of health behaviors, in predicting incidence of hypertension during later childhood and adolescence.

## Methods

### Sample

The FNPA was implemented as standard of care for pediatric well-child visits in a large mid-Atlantic health system beginning in 2013. Data have been collected from young children (ages 3- 9 years) from 2013-2019 and the age range was expanded to 12 years in 2020. The data used in this study were drawn from a larger Geisinger Institutional Review Board (IRB) approved database (IRB # 2023-1710). Clinical electronic health record and demographic data for the present study were obtained from annual well-child visits conducted between the years 2013 to 2022, resulting in a total of 51,691 children associated with 1,075,200 individual visit records. To be included in the present analysis, the sample was limited to participants who met the following criteria for the baseline period: 1) age between 3 and 9 years on or after 2013, 2) at least two records of BMI at separate visits to evaluate overweight status, 3) at least one record of FNPA completion, and 4) no history of hypertension diagnosis or elevated blood pressure (required a minimum of 2 separate blood pressure readings). For the follow-up period, 5) age between 9 and 18 years on or before 2022, and 6) at least three separate visits with blood pressure measurements recorded. The baseline period captures both early and middle childhood and maximizes the available sample for follow-up.

### Measures

#### FNPA

The FNPA risk assessment is digitally completed by parents or caregivers in the patient portal or on an electronic device in the waiting room, in conjunction with annual well-child visits during the participant’s baseline period. When annual well-child visits during the baseline period resulted in multiple FNPA assessments, these were averaged to obtain a baseline score. The FNPA data were coded to produce the FNPA summary score (Table 1).^10–11^ For the present analyses that explores the potential moderating influence of FNPA on incident hypertension, each participant was categorized into one of 3 FNPA categories using the 25th and 75th percentiles (61.5 and 70, respectively) of the FNPA summary score: scores less than the 25^th^ percentile were categorized as ‘low’, values between 25^th^ and 75^th^ percentile as ‘moderate’ and values greater than the 75^th^ percentile as ‘high’. This approach uses the observed data and mirrors the established pattern from prior studies as lower FNPA summary scores have been associated with obesity risk and higher scores have been associated with protection against obesity.^10–14^

### BMI Change Indicator

Overweight status and change in overweight status was defined using the BMI85, also known as % overweight, and is the scaled distance away from the BMI value at the age- and sex-specific 85th percentile ^9^. Specifically, BMI85 is calculated as

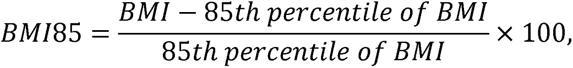

where the 85th percentile is acquired from CDC growth charts for males and females from 2 to 20 years old. To evaluate change in overweight status during childhood, each participant was categorized into one of the BMI85 groups (loss, stable, or gain) based on the 25th and 75th percentiles (-1.95 and 1.41, respectively) of the annualized BMI85 change score (ΔBMI85), which was calculated from their first and last BMI85 records during the baseline period.

Overweight status reflects change scores that were less than the 25^th^ percentile were categorized as ‘loss’, values between 25^th^ and 75^th^ percentile as ‘stable’ and values greater than the 75^th^ percentile as ‘gain’. The discretization of annual BMI85 changes provided robustness against the influence of outliers due to potential short time interval and measurement error of BMI records within the specified age range.

### Hypertension Status

The primary outcome was defined as a binary variable denoting the presence or absence of blood pressure readings ≥95th percentile (e.g., Stage 1 or Stage 2 HTN) at a minimum of three separate visits during the follow-up period. The data to define the variable originated from electronic health records that were evaluated for blood pressure measurements beyond the age of 9 years. For children aged 9 to 13 years, blood pressure percentiles were determined based on age, height, and sex, per American Academy of Pediatrics guidance.^17^ Blood pressure values were categorized as normal (50^th^ percentile); elevated (>90^th^ percentile); stage 1 HTN (≥ 95^th^ percentile); and stage 2 HTN (≥95^th^ percentile + 12 mm Hg). For children aged 13 years or older, thresholds followed guidance from the American Heart Association and American College of Cardiology for HTN in adults.^18^ For participants with elevated blood pressure, stage 1 HTN, or stage 2 HTN during the follow-up period, we defined the age at which hypertension was first discovered after reaching 9 years old as the event time. Participants without hypertension were included in the analysis until the end of the follow-up period at the age of 18 years.

### Analyses

We evaluated the empirical rate of hypertension for each FNPA score and BMI85 change group, namely the proportion of participants having ≥3 elevated blood pressure measures during the follow-up period, and we conducted log-rank tests to examine the difference across groups, controlling for baseline BMI. Cox proportional hazards models were performed to assess the risk of hypertension development, with the reference groups denoted as the moderate FNPA score group and the BMI85 stable group. In addition to categorical FNPA score and BMI85 change divisions as baseline characteristics, potential confounding variables were incorporated in the model, including sex, race and ethnicity, and insurance type. Hazard ratios (HRs) and 95% confidence intervals (CIs) relating BMI85 change and FNPA score were estimated from the model. The assumption of time-invariant hazard ratios and the significance of explanatory variables was tested. Kaplan-Meier curves were used to compare the empirical rate of hypertension development for different subgroups.

## Results

Participants were classified into subgroups in a 3 by 3 manner, i.e., FNPA low, moderate, high and BMI85 change status loss, stable, and gain during the baseline period. Participant characteristics are described by FNPA summary scores and change in overweight status during the baseline period in Table 2. Among the study sample, a majority were female (n= 2,060, 54.1%), white (n=3,127, 82.1%), and publicly insured (n=2,192, 57.6%).

**Table 2.** Characteristics of children during the baseline period by Family Nutrition and Physical Activity (FNPA) summary score category and overweight status.

| Characteristic | Category | Sample<br>(N=3808)<br>N(%) | FNPA<br>Low<br>N(%) | FNPA<br>Moderate<br>N(%) | FNPA<br>High<br>N(%) | Overweight<br>Loss<br>N(%) | Overweight<br>Stable<br>N(%) | Overweight<br>Gain<br>N(%) |
| --- | --- | --- | --- | --- | --- | --- | --- | --- |
| Sex | Female | 2060 (54.1) | 490 (51.4) | 1135 (54.6) | 435 (56.1) | 528 (55.5) | 992 (52.1) | 540 (56.7) |
|  | Male | 1748 (45.9) | 464 (48.6) | 944 (45.4) | 340 (43.9) | 424 (44.5) | 912 (47.9) | 412 (43.3) |
| Race/<br>ethnicity | White | 3127 (82.1) | 727 (76.2) | 1744 (84.0) | 656 (84.7) | 790 (83.0) | 1610 (52.1) | 727 (76.3) |
|  | Black | 256 (5.7) | 99 (10.4) | 123 (5.9) | 34 (4.4) | 54 (5.7) | 111 (5.8) | 91 (9.6) |
|  | Hispanic<br>or Latino | 356 (9.4) | 112 (11.7) | 173 (8.3) | 70 (9.0) | 95 (10.0) | 145 (7.6) | 116 (12.2) |
|  | Other | 69 (1.3) | 16 (1.7) | 38 (1.8) | 15 (1.9) | 13 (1.3) | 38 (2.0) | 18 (1.9) |
| Insurance | Public | 2192 (57.6) | 664 (69.6) | 1177 (56.6) | 351 (45.3) | 549 (57.7) | 1065 (55.9) | 578 (60.7) |
|  | Private | 1615 (42.4) | 290 (30.4) | 902 (43.4) | 424 (54.7) | 403 (42.3) | 839 (44.1) | 374 (39.3) |

Table 3 presents the rate of hypertension for each group and the average age at the initial discovery of hypertension, as indicated by 3 separate blood pressure readings. Children who gained BMI85 during the baseline period exhibited a notably higher incidence of hypertension development (41.7%) compared to those that were overweight stable (29.9%) and the overweight loss group (27.7%). The differences in the incidence of hypertension were found significant among the BMI85 change groups (P < 0.001), whereas it was insignificant among the FNPA score groups (P = 0.600).

**Table 3.** Descriptive characteristics regarding hypertension (HTN) over the 6-year follow-up period by baseline Family Nutrition and Physical Activity (FNPA) summary score and overweight status.

|  | FNPA<br>Low | FNPA<br>Moderate | FNPA<br>High | Overweight<br>Loss | Overweight<br>Stable | Overweight<br>Gain |
| --- | --- | --- | --- | --- | --- | --- |
| **Rate of HTN\*** | 35.2% | 32.1% | 29.3% | 27.3% | 29.9% | 41.7% |
| **Age at initial HTN**<br>**(Mean, SD)** | 11.3 (1.9)<br>years | 11.2 (1.9)<br>years | 11.2 (1.8)<br>years | 11.2 (1.9)<br>years | 11.3 (1.9)<br>years | 11.0 (1.7)<br>years |

\*HTN was defined using blood pressure measures obtained at three separate visits and interpreted with age-specific guidelines

Table 4 summarizes the model results. The time-invariant hazard assumption was met for the model (P = 0.172). Model results revealed a significantly lower risk of hypertension among Hispanic or Latino vs. white participants (HR 0.80; 95% CI 0.66-0.98), but no sex differences were observed. Publicly insured participants had a significant 1.3 times increased risk compared to those with private insurance (HR 1.30; 95% CI 1.15-1.48). Additionally, likelihood ratio tests revealed that both overweight status (P < 0.001) and FNPA score (P = 0.048), as well as their interactions (P = 0.037) demonstrated statistically significant explanatory power regarding the incidence of hypertension. After adjusting for all other factors and compared to the reference overweight stable group, participants in the overweight gain group were at nearly twofold increased risk of hypertension development (HR 2.01; 95% CI, 1.65-2.44), whereas those in the overweight loss group were associated with a 0.75 times reduced risk (HR 0.75; 95% CI, 0.59- 0.96).

**Table 4.** Estimated hypertension coefficients and hazard ratios (95% confidence interval) of the Cox proportional hazards model.

| | | Coefficient $\beta$ | Hazard ratio<br>$exp(\beta)$ | p-value |
| --- | --- | --- | --- | --- |
| Overweight<br>Status | Loss | -0.28 | 0.75 (0.59-0.96) | 0.02 |
|  | Stable | Reference |  |  |
|  | Gain | 0.70 | 2.01 (1.65-2.44) | <0.001 |
| FNPA score | Low | 0.07 | 1.07 (0.87-1.32) | 0.52 |
|  | Moderate | Reference |  |  |
|  | High | 0.19 | 1.21 (0.98-1.50) | 0.08 |
| Sex | Male | Reference |  |  |
|  | Female | -0.07 | 0.93 (0.83-1.04) | 0.22 |
| Race and<br>ethnicity | White | Reference |  |  |
|  | Black | 0.03 | 1.05 (0.82-1.30) | 0.78 |
|  | Hispanic or Latino | -0.22 | 0.80 (0.66-0.98) | 0.03 |
|  | Others | -0.01 | 0.99 (0.59-1.65) | 0.96 |
| Insurance | Private | Reference |  |  |
|  | Public | 0.26 | 1.30 (1.15-1.48) | <0.001 |
| Interactions | FNPA: low BMI85: loss | 0.22 | 1.25 (0.87-1.81) | 0.23 |
|  | FNPA: high BMI85: loss | 0.17 | 1.19 (0.81-1.73) | 0.38 |
|  | FNPA: low BMI85: gain | -0.08 | 0.92 (0.68-1.26) | 0.61 |
|  | FNPA: high BMI85: gain | -0.46 | 0.63 (0.44-0.91) | 0.01 |

The interaction term between high FNPA score and BMI85 gain was significant (HR 0.63; 95% CI, 0.44-0.91) and suggests a potential relationship between high FNPA scores (healthy behaviors) and lower rates of hypertension development within the BMI85 gain group. Figure 1 illustrates the estimated hazard ratios and corresponding 95% CIs for each of the 3 by 3 subgroups. No significant differences were found among subgroups without BMI85 gain.

**Figure 1.**
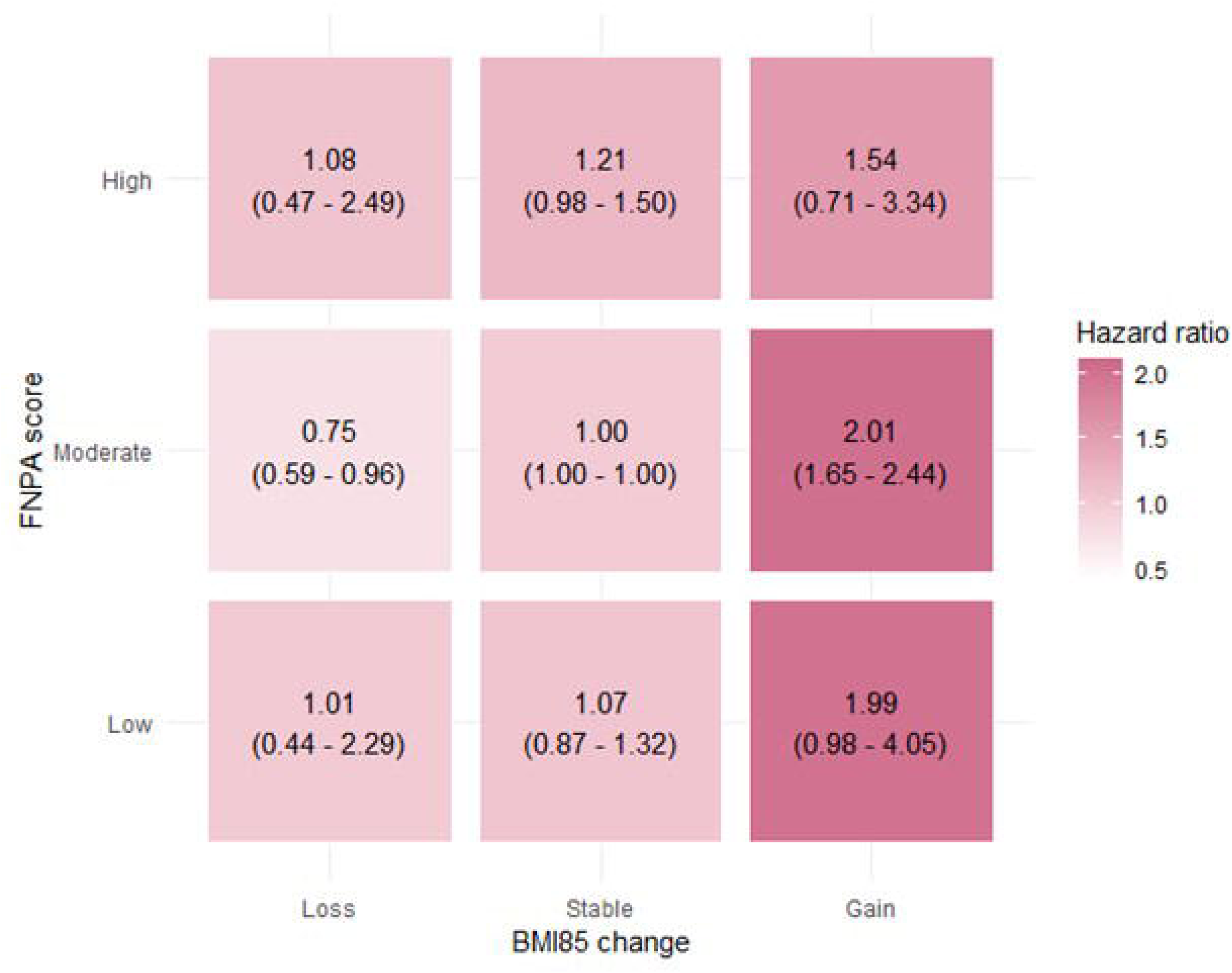
Estimated hazard ratios (95% confidence intervals) of hypertension based on BMI85 change and FNPA score subgroups.

Specifically, children who gained BMI85 during the baseline period had a higher risk of hypertension compared to those who maintained or lost BMI85 from ages 3 to 9 years. Among children with moderate or low FNPA scores, those who experienced BMI85 gain had a nearly twofold increased hazard of hypertension compared to those who were BMI85 stable. Among children gaining overweight, high FNPA scores were associated with 25% lower risk of hypertension compared to those with moderate or low FNPA scores.

Figure 2 presents the cumulative survival probability of hypertension during the follow-up period by Kaplan-Meier curves. The BMI85 gain group demonstrated lower survival probabilities of hypertension compared to the other two BMI85 status groups across all three levels of FNPA score, which corresponded to the results of the proportional hazards model.

**Figure 2.**
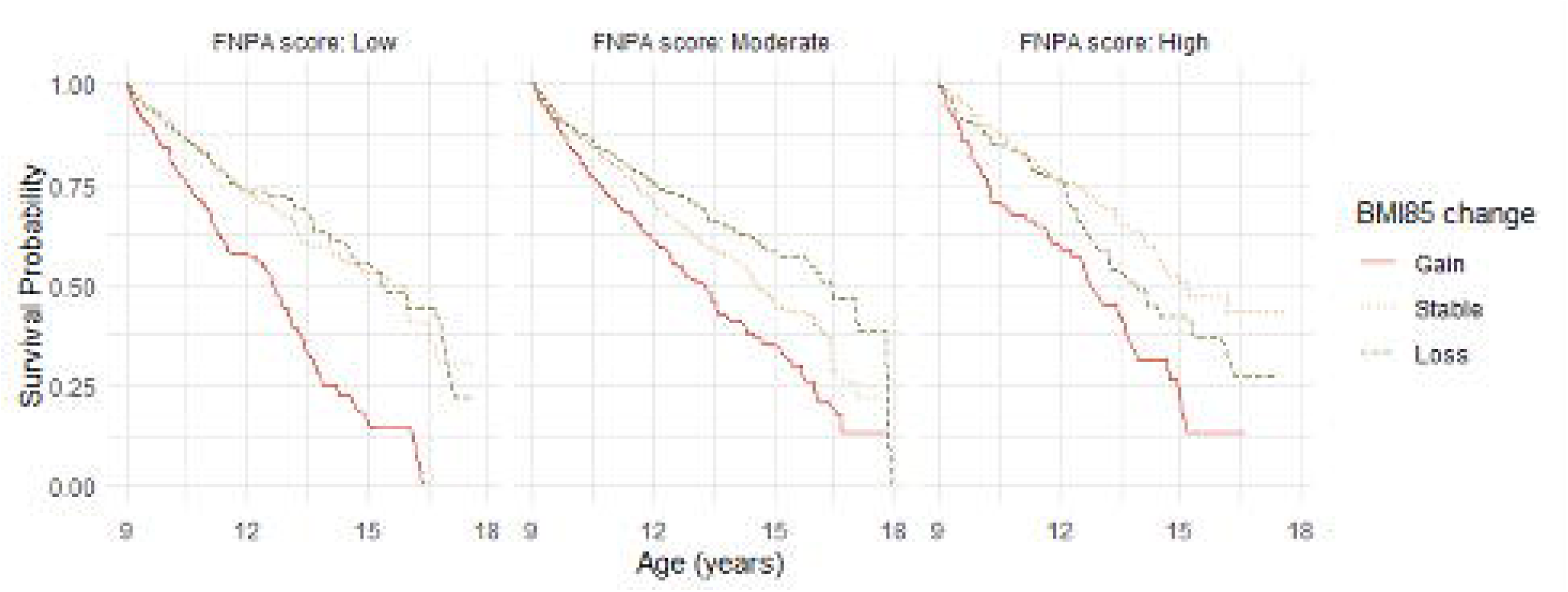
Kaplan-Meier survival curves of hypertension after the age of 9 years.

Specifically, this discrepancy was most pronounced at the low FNPA score level indicating unhealthy behaviors, highlighting the lower risk of hypertension associated with higher FNPA scores indicating healthy behaviors.

## Discussion

Our main finding is that behaviors associated with the FNPA summary score attenuated the incidence of hypertension associated with BMI85 gain among children with overweight. This suggests that FNPA screening at well-child visits and preventative counseling to promote healthy home environments offers protection from the early onset of hypertension associated with increasing overweight among children who were not hypertensive at baseline. These findings complement prior research that demonstrates the utility of FNPA screening and counseling on the primary prevention of childhood obesity^13^ and its potential as a novel surrogate measure for cardiovascular disease in childhood ^19^

The FNPA was developed and validated to assess parenting practices, child behaviors, and home environment characteristics that may predispose children to obesity.^9–12^ The FNPA screening tool addresses the home environment and behaviors relevant to effective obesity prevention including 1) poor diet (e.g., consumption of sugar-sweetened beverages and energy-dense foods); 2) low levels of physical activity; 3) short sleep duration; 4) sedentary behaviors (e.g., high media use); and 5) parenting practices in the home environment. When delivered as standard of care during annual well-child visits and with clinical decision support, FNPA results enable pediatricians to quickly assess a child’s risk and provide personalized preventative counseling to the child and their family.^20^ Higher FNPA summary scores are associated with healthier weight status, as a 1-point increment in score has been associated with a 0.12 lower BMI.^14^ This offers clinicians and families a numerical benchmark for tracking prevention and treatment progress.^13,14^

Further, this study informs identified research gaps in the obesity clinical practice guidelines related to the age to begin evaluation for cardiometabolic comorbidities.^17^ Our findings suggest that hypertension diagnoses emerged as early as age 10 years. Consistent with prior studies, our findings support the direct association between childhood overweight and obesity and hypertension development. Our threshold for evaluating the association was set at the 85^th^ percentile of BMI-for-age and -sex, however this has also been observed at the 60^th^ percentile.^21^ The proportion of children with overweight during early childhood who developed hypertension during latter childhood and adolescence was higher in our population than what others have reported.^2,3^ Given pediatricians care for children over an extended period of time, we call attention to the pattern of continued body mass gain among children with overweight, as 4 in 10 developed hypertension. Our findings add to the evidence base regarding the benefits of screening for high blood pressure in youth aged 3 to 18 years, currently evaluated as insufficient per the USPSTF.^22^ This study by consequence, also informs an identified USPSTF research gap by examining the associations of childhood hypertension and surrogate measures of cardiovascular disease, namely the FNPA score, overweight and obesity in childhood.

The findings were evaluated using change in BMI85 which provide advantages over traditional BMI indicators for examining longitudinal changes in weight status.^23–25^ For those participants with relatively high BMI, BMI85 also outperforms the original BMI z-score or BMI percentile in addressing the limitation from the right skewed nature of the BMI distribution conditional on age.^10^

The results presented in this analysis underscore the value of the FNPA as a screening tool and in identifying positive behaviors. The lower incidence of hypertension in those with unhealthy FNPA scores suggests behavioral intervention in those with poor scores may have health benefits beyond obesity prevention. The FNPA has been increasingly used in clinical settings to facilitate implementation of anticipatory guidance and is available in the Epic^®^ electronic health record.

The primary prevention of childhood obesity is markedly more effective than secondary or tertiary care for obesity co-morbidities. Given the near-universal exposure to well-child visits for children in the United States, pediatric providers may be supported by parent responses to FNPA to identify modifiable risk factors, inform preventive counseling and treatment, and thereby better meet the urgent need to prevent and reduce childhood obesity and its downstream effects including hypertension.

### Strengths & Limitations

A distinct strength of this study is the use of a large clinical data set with clinical measures of blood pressure and BMI, as well as parent-reported FNPA data. We recognize the potential limitation in assuming that a single FNPA assessment provides a stable indicator of home environment; however, it is relevant that this single indicator of risk has predictive utility over this timespan. The retrospective design is a further limitation. Future studies may investigate the stability of the FNPA during youth development as well as the relative importance of different behavioral constructs. By convenience, our study was limited to FNPA exposures during the preschool and early elementary years. Future studies may examine the associations between FNPA, weight status, and hypertension among older children and adolescents. Finally, the observational study design allows for investigation of associations but not causality. Clinical trials are needed to examine the outcomes of behavioral intervention based on FNPA summary scores and incident hypertension among children with overweight to confirm the observations in this study.

## Conclusion

The prevention of childhood obesity and its comorbidities including hypertension can be facilitated through effective screening tools like FNPA that are delivered via pediatric care settings. We found that healthy behaviors assessed with the FNPA among children who became more overweight during childhood were potentially linked to a decreased risk of developing hypertension during latter childhood and adolescence. Efforts must continue to be made to implement and learn from tools like the FNPA that promote healthy home environments to better prevent and treat childhood obesity and its downstream effects on cardiometabolic health.

## Data Availability

The datasets generated and/or analyzed during the current study are not publicly available due to being protected electronic health record data.

## Contributors Statement Page

Drs. Lisa Bailey-Davis and Gregory Welk conceptualized and designed the study, drafted the initial manuscript, and critically reviewed and revised the manuscript.

Dr. Carolyn McCabe drafted the initial manuscript, and critically reviewed and revised the manuscript.

Chengpeng Zeng designed the carried out the initial analyses, and critically reviewed and revised the manuscript.

Dr. Franceschelli Hosterman, Adam Cook, and Craig Wood carried out data collection and critically reviewed and revised the manuscript.

Drs. Shuping Ge and Samuel Gidding critically reviewed and revised the manuscript.

All authors approved the final manuscript as submitted and agree to be accountable for all aspects of the work.

## Acknowledgements

The authors acknowledge the contributions of the FNPA working group (myfnpa.org) and their feedback on early findings and support for promoting the availability of FNPA in the Epic^®^ Electronic Health Record.

## References

1. Song P, Zhang Y, Yu J, et al. Global Prevalence of Hypertension in Children: A Systematic Review and Meta-analysis. JAMA Pediatr. 2019 Dec 1;173(12):1154–1163.

2. Tu W, Eckert GJ, DiMeglio LA, Yu Z, Jung J, Pratt JH. Intensified effect of adiposity on blood pressure in overweight and obese children. Hypertension. 2011 Nov;58(5):818–24.

3. Sorof JM, Lai D, Turner J, Poffenbarger T, Portman RJ. Overweight, Ethnicity, and the Prevalence of Hypertension in School-Aged Children. Pediatrics March 2004; 113 (3): 475–482. 10.1542/peds.113.3.475

4. de Simone G, Devereux RB, Izzo R, et al. Lack of reduction of left ventricular mass in treated hypertension: the strong heart study. J Am Heart Assoc. 2013 Jun 6;2(3):e000144.

5. Carullo N, Zicarelli M, Michael A, et al. Childhood Obesity: Insight into Kidney Involvement. Int J Mol Sci. 2023 Dec 12;24(24):17400.

6. Myette RL, Flynn JT. The ongoing impact of obesity on childhood hypertension. Pediatr Nephrol. 2024 Aug;39(8):2337–2346.

7. Hampl SE, Hassink SG, Skinner AC, et al. Clinical Practice Guideline for the Evaluation and Treatment of Children and Adolescents with Obesity. Pediatrics (Evanston). 2023;151(2):1.

8. Grossman DC, Bibbins-Domingo K, Curry SJ, et al. Screening for obesity in children and adolescents: US preventive services task force recommendation statement. JAMA. 2017;317(23):2417–2426.

9. Peyer KL, Bailey-Davis L, Welk G. Development, applications, and refinement of the family nutrition and physical activity (FNPA) child obesity prevention screening. Health Promot Pract. 2021;22(4):456–461.

10. Peyer KL, Welk GJ. Construct validity of an obesity risk screening tool in two age groups. International Journal of Environmental Research and Public Health. 2017;14(4):419.

11. Ihmels MA, Welk GJ, Eisenmann JC, Nusser SM. Development and preliminary validation of a Family Nutrition and Physical Activity (FNPA) screening tool. Int J Behav Nutr Phys Act. 2009 Mar 12;6:14.

12. Ihmels MA, Welk GJ, Eisenmann JC, Nusser SM, Myers EF. Prediction of BMI change in young children with the family nutrition and physical activity (FNPA) screening tool. Ann Behav Med. 2009;38(1):60–68.

13. Bailey Davis L, Kling SMR, Wood GC, et al. Feasibility of enhancing well child visits with family nutrition and physical activity risk assessment on body mass index. Obes Sci & Pract. 2019;5(3):220–230.

14. Tucker JM, Howard K, Guseman EH, Yee KE, Saturley H, Eisenmann JC. Association between the Family Nutrition and Physical Activity Screening Tool and obesity severity in youth referred to weight management. Obes Res Clin Pract. 2017 May-Jun;11(3):268–275.

15. Lal JC, Margai L, Zitkovsky HS, Price LL, González S. Improving Health Behaviors and Weight Parameters With Motivational Interviewing and the TEEEN Program in an Ethnically and Socioeconomically Diverse Pediatric Population. American Journal of Medicine Open. 2023 Dec 1;10:100042.

16. Helsel BC, Foster RNS, Sherman J, Steele R, Ptomey LT, Montgomery R, Washburn RA, Donnelly JE. The Family Nutrition and Physical Activity Survey: Comparisons with Obesity and Physical Activity in Adolescents with Autism Spectrum Disorder. J Autism Dev Disord. 2023 Jan;53(1):89–95.

17. Flynn JT, Kaelber DC, Baker-Smith CM, et al. SUBCOMMITTEE ON SCREENING AND MANAGEMENT OF HIGH BLOOD PRESSURE IN CHILDREN. Clinical Practice Guideline for Screening and Management of High Blood Pressure in Children and Adolescents. Pediatrics. 2017 Sep;140(3):e20171904.

18. Whelton PK, Carey RM, Aronow WS, et al. 2017 ACC/AHA/AAPA/ABC/ACPM/AGS/APhA/ASH/ASPC/NMA/PCNA Guideline for the Prevention, Detection, Evaluation, and Management of High Blood Pressure in Adults: Executive Summary: A Report of the American College of Cardiology/American Heart Association Task Force on Clinical Practice Guidelines. Hypertension. 2018 Jun;71(6):1269–1324.

19. Yee KE, Eisenmann JC, Carlson JJ, Pfeiffer KA. Association between The Family Nutrition and Physical Activity Screening Tool and cardiovascular disease risk factors in 10-year old children. Int J Pediatr Obes. 2011 Aug;6(3-4):314–20.

20. Christison AL, Daley BM, Asche CV, Ren J, Aldag JC, Ariza AJ, Lowry KW. Pairing motivational interviewing with a nutrition and physical activity assessment and counseling tool in pediatric clinical practice: a pilot study. Child Obes. 2014 Oct;10(5):432–41. doi: 10.1089/chi.2014.0057.

21. Koebnick C, Sidell MA, Li X, Woolford SJ, Kuizon BD, Kunani P. Association of High Normal Body Weight in Youths With Risk of Hypertension. JAMA Netw Open. 2023;6(3):e231987.

22. US Preventive Services Task Force; Krist AH, Davidson KW, Mangione CM, et al. Screening for High Blood Pressure in Children and Adolescents: US Preventive Services Task Force Recommendation Statement. JAMA. 2020 Nov 10;324(18):1878–1883.

23. Rodig NM, Roem J, Schneider MF, Seo-Mayer PW, Reidy KJ, Kaskel FJ, Kogon AJ, Furth SL, Warady BA. Longitudinal outcomes of body mass index in overweight and obese children with chronic kidney disease. Pediatr Nephrol. 2021 Jul;36(7):1851–1860.

24. Kerr JA, Dumuid D, Downes M, Lange K, O’Connor M, Thornton L, Mavoa S, Lycett K, Olds TS, Edwards B, O’Sullivan JM, Juonala M, Burgner D, Wake M. Socioeconomic disadvantage and polygenic risk for high BMI magnify obesity risk across childhood: a longitudinal, population, cohort study. Lancet Glob Health. 2023 Mar;11 Suppl 1:S9–S10.

25. Burke NJ, Hellman JL, Scott BG, Weems CF, Carrion VG. The impact of adverse childhood experiences on an urban pediatric population. Child Abuse Negl. 2011 Jun;35(6):408–13.

